# Biallelic GTF3A mutations underline a novel human combined immunodeficiency

**DOI:** 10.64898/2026.07.24.26358408

**Authors:** Lang Yu, Hailin Li, Qinglv Wei, Jijun Wu, Yulin Li, Liwen Zhang, Wenhui Li, Lina Zhou, Yanjun Jia, Ying Dou, Qing Zhou, Xiaodong Zhao, Yunfei An

**Author notes:** Corresponding Authors: Yunfei An, Department of Rheumatology and Immunology, Children’s Hospital of Chongqing Medical University, No. 136, Zhongshan 2nd Road, Yuzhong District, Chongqing 400014, China;, Xiaodong Zhao, Department of Rheumatology and Immunology, Children’s Hospital of Chongqing, Medical University, No. 136, Zhongshan 2nd Road, Yuzhong District, Chongqing 400014, China. Lang Yu, Hailin Li, QingLv Wei and Jijun Wu contributed equally to this study and are co-first authors. Funding: Chongqing Natural Science Foundation Innovation and Development Joint Fund (CSTB2024NSCOLZX0100), Chongqing medical scientific research project NO.2024GGXM004 (Joint project of Chongqing Health Commission and Science and Technology Bureau), Clinical Research Project for the Summit Program of Children’s Hospital of Chongqing Medical University (CHCMU-2024-XKDF-1001).

## Abstract

Ribosome biogenesis defects are increasingly recognized in hematologic disorders, yet their contribution to human combined immunodeficiency (CID) remains largely unexplored. Here, we identify compound heterozygous mutations in GTF3A, encoding transcription factor IIIA (TFIIIA), in a patient with CID presenting with profound T-cell lymphopenia, diminished thymic output, and humoral failure. The patient-derived TFIIIA variants, I267S and L364Yfs*37, disrupted 5S rRNA transcription, impaired RNA-binding capacity, and compromised protein stability. Patient T and B lymphocytes exhibited intrinsic proliferative and differentiation defects, as well as increased apoptotic susceptibility in T cells, recapitulating the clinical phenotype. To model TFIIIA dosage, we generated heterozygous and progressively depleted Jurkat cell clones via sequential CRISPR editing; complete TFIIIA loss was lethal, whereas graded reduction impaired proliferation in a dose-dependent manner, fully rescued by wild-type GTF3A reconstitution. Collectively, these findings establish deficiency of GTF3A resulting in combined immunodeficiency (DoGCID) within the spectrum of ribosomopathies, highlighting the exquisite sensitivity of lymphocyte fitness to disruptions in ribosome biogenesis.

**One sentence summary:** Compound heterozygous GTF3A mutations impair TFIIIA-dependent 5S rRNA transcription and ribosome biogenesis, causing intrinsic T and B cell proliferative defects that manifest as combined immunodeficiency in humans.

## Introduction

The molecular dissection of human immunodeficiencies has historically relied on the identification of monogenic lesions that disrupt lymphocyte development or function. Among these, combined immunodeficiencies (CIDs) are defined by partial yet clinically significant defects in T cell immunity, frequently accompanied by humoral abnormalities affecting B cells and antibody production, while natural killer (NK) cell compartments may also be variably involved (*1, 2*). The clinical spectrum of CID is notably broad—ranging from late-onset opportunistic infections to severe immune dysregulation—which often delays diagnosis and obscures the underlying genetic etiology. Despite the widespread application of next-generation sequencing, a substantial fraction of CID patients, estimated at 30–40%, remain without a molecular diagnosis, underscoring the existence of unrecognized genetic determinants that govern T cell homeostasis (*3, 4*).

Ribosome biogenesis represents an essential yet largely unexplored frontier in the pathogenesis of CID. While global disruption of protein synthesis is expected to be incompatible with life, partial or tissue-restricted defects in ribosomal components have emerged as a distinct class of human diseases—ribosomopathies—that frequently manifest with cell-type-specific phenotypes, including hematologic failure and immune deficiency (*5, 6*). Among the myriad factors controlling ribosome assembly, transcription factor IIIA (TFIIIA), encoded by GTF3A, occupies a unique position. Unlike general transcription factors, TFIIIA is dedicated exclusively to the synthesis of 5S rRNA, an integral scaffold of the 60S large ribosomal subunit (*7*). This 42-kDa zinc-finger protein executes a dual function: it initiates RNA polymerase III–dependent transcription at the 5S rDNA internal control region, and concurrently chaperones the nascent 5S rRNA transcript toward proper ribosome incorporation (*8–10*). The centrality of 5S rRNA to ribosome function would predict that TFIIIA dysfunction carries profound consequences; yet, human diseases arising from GTF3A mutations have remained conspicuously absent from the literature—until a recent report linked biallelic GTF3A variants to susceptibility to herpes simplex virus encephalitis, accompanied by CVID-like humoral defects (*11*). That study hinted at broader lymphoid perturbations, but whether TFIIIA deficiency can intrinsically disrupt T cell development in humans, and whether such disruption suffices to produce a bona fide CID phenotype, has not been established.

Here, we report the identification of compound heterozygous GTF3A mutations in a CID patient presenting with profound T cell lymphopenia, diminished thymic output, recurrent infections, and concomitant B cell abnormalities. Using patient-derived primary cells and CRISPR-engineered cellular models, we demonstrate that TFIIIA loss disrupts 5S rRNA biogenesis, leading to impaired proliferation and enhanced apoptotic susceptibility in lymphocytes. Our findings establish deficiency of GTF3A resulting in combined immunodeficiency (DoGCID) within the spectrum of ribosomopathies.

## Results

### Clinical course of the patient with CID

The patient was hospitalized in the neonatal period for pneumonia and improved with antibiotic therapy. During infancy, he developed recurrent infections, diarrhea, and failure to thrive. Subsequently, the patient was hospitalized for severe pneumonia, with chest CT revealing diffuse bilateral pulmonary infiltrates **(Fig. 1A)**. Bronchoalveolar lavage fluid tested positive for cytomegalovirus (CMV) DNA and silver staining, suggestive of CMV pneumonia with Pneumocystis jirovecii co-infection. Laboratory findings showed leukopenia and lymphopenia **(Fig. 1B)**. Immunological evaluation revealed profound CD3⁺ T-cell lymphopenia (622.3 cells/µL), a reduced CD4/CD8 ratio (0.72), reduced B-cell counts and normal NK cell counts **(Table 1)**, and pan-hypogammaglobulinemia with markedly reduced IgG, IgM, and IgA levels **(Fig. 1C)**, consistent with a diagnosis of CID. Monthly intravenous immunoglobulin (IVIG) replacement therapy was initiated at 5 g per dose, later adjusted to 400–600 mg/kg, resulting in normalization of serum IgG levels. Trimethoprim-sulfamethoxazole (TMP-SMX) prophylaxis for P. jirovecii was also administered.

**Figure 1.**
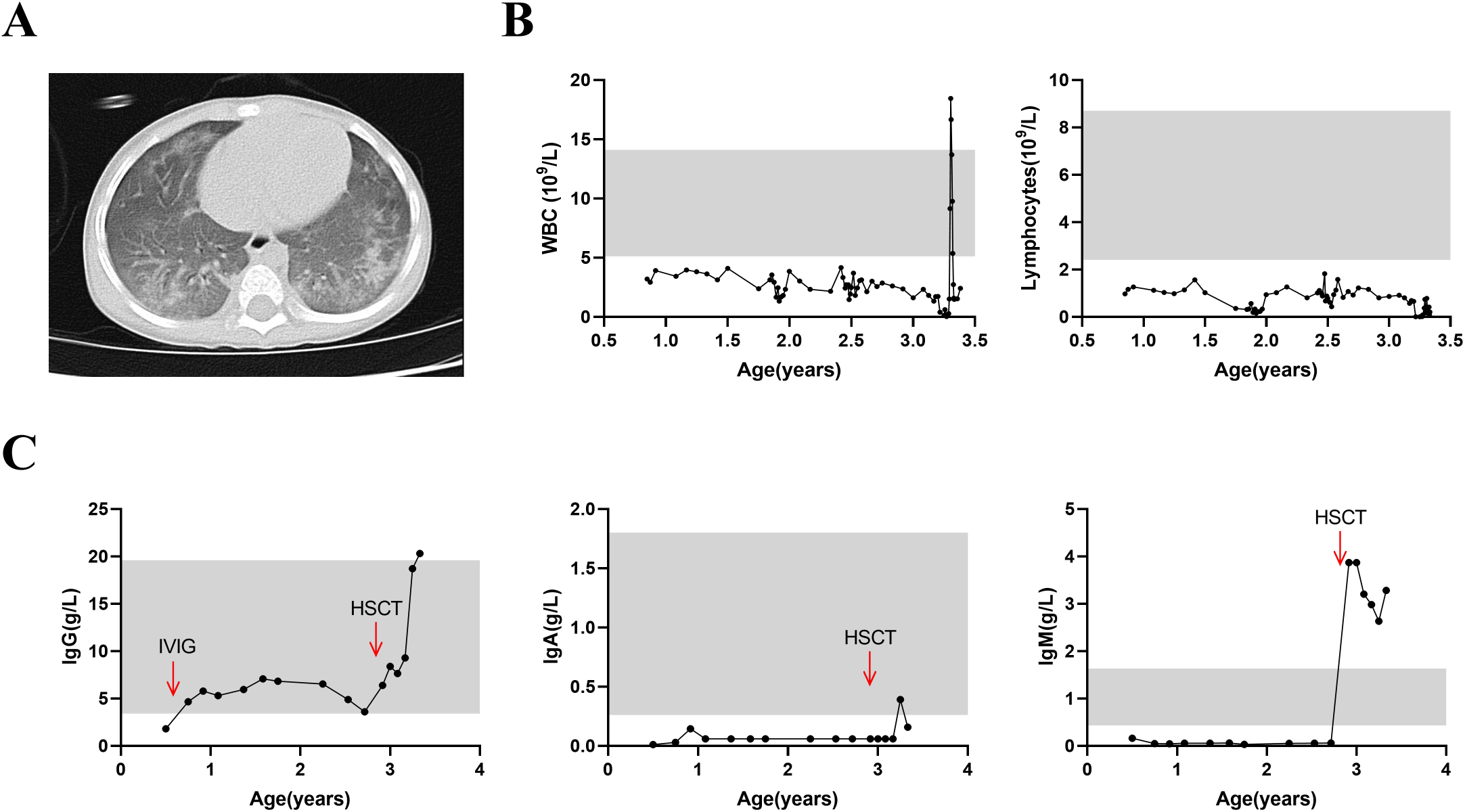
Clinical characteristics of the patient with biallelic GTF3A mutations. (A) Chest CT scan showing extensive bilateral pulmonary infiltrates due to cytomegalovirus (CMV) and Pneumocystis jirovecii (PCP) co-infection. (B) Absolute counts of peripheral blood leukocytes and lymphocytes over time. (C) Longitudinal serum levels of immunoglobulins IgG, IgM, and IgA. Red arrows indicate monthly intravenous immunoglobulin (IVIG) infusions and hematopoietic stem cell transplantation (HSCT) .

**Table 1.** Immunological Characteristics of the Patient.

|  | 6-9months |  |  |  | 1-3years |  |  |  |
| --- | --- | --- | --- | --- | --- | --- | --- | --- |
|  | Proportion | Proportion Reference | Absolute count | Absolute count Reference | Proportion | Proportion Reference | Absolute count | Absolute count Reference |
| CD3+ cells | 29.9 | 55.32-73.11 | 622.3 | 2187-6352 | 43.2 | 53.88-72.87 | 380.4 | 1794-4247 |
| CD3+CD8+ cells | 15.4 | 15.88-31.48 | 319.7 | 686-2278 | 21.8 | 19.00-32.51 | 191.5 | 580-1735 |
| CD3+CD8+CD27+CD45RA+ cells (Naive) | 39.5 | 47.36-92.45 | 126.3 | 535-1677 | 37.9 | 36.80-83.16 | 72.6 | 356-1095 |
| CD3+CD8+CD27+CD45RA- cells (CM) | 45.3 | 1.0-24.2 | 144.8 | 51-316 | 27.7 | 5.18-31.66 | 53.0 | 56-406 |
| CD3+CD8+CD27-CD45RA- cells (EM) | 4.0 | 0.0-5.8 | 12.8 | 2-120 | 9.4 | 0.70-11.22 | 18.1 | 6-145 |
| CD3+CD8+CD27-CD45RA+ cells (TEMRA) | 11.1 | 0-15.2 | 35.5 | 2-430 | 24.9 | 0.84-33.02 | 47.7 | 9-440 |
| CD3+CD4+ cells | 11.1 | 28.17-47.74 | 231.1 | 1125-3768 | 12.1 | 24.08-42.52 | 106.6 | 902-2253 |
| CD3+CD4+CD27+CD45RA+ cells (Naive) | 65.3 | 59.28-88.09 | 150.9 | 764-2972 | 38.8 | 46.14-84.40 | 41.4 | 472-1760 |
| CD3+CD4+CD27+CD45RA- cells (CM) | 28.0 | 10.15-33.38 | 64.70 | 206-796 | 49.3 | 13.88-48.12 | 52.5 | 212-735 |
| CD3+CD4+CD27-CD45RA- cells (EM) | 5.8 | 0.1-1.5 | 13.4 | 11-60 | 11.4 | 0.94-6.46 | 12.2 | 15-87 |
| CD3+CD4+CD27-CD45RA+ cells (TEMRA) | 1.0 | 0-0.8 | 2.2 | 0-33 | 0.5 | 0.00-1.36 | 0.5 | 0-22 |
| CD4/CD8 | 0.72 | 0.93-2.52 |  |  | 0.60 | 0.9-2.13 |  |  |
| CD19+ cells | 36.7 | 17.20-29.71 | 764.2 | 916-1832 | 50.1 | 13.23-26.39 | 440.4 | 461-1456 |
| CD3-CD56+ cells | 33.3 | 5.67-15.90 | 693.5 | 306-896 | 6.7 | 7.21-20.90 | 58.8 | 270-1053 |
| TCRαβ+DNT cells | 0.4 | 0.41-1.55 | 2.6 | 16-58 | 0.7 | 0.37-1.80 | 2.7 | 9-57 |
| γδ T cells | 21.8 | 3.95-10.40 | 135.7 | 128-436 | 31.1 | 4.94-17.98 | 118.3 | 114-539 |
| TRECs (copies/ul blood) |  |  | 15 | >40 |  |  |  |  |
| KRECs (copies/ul blood) |  |  | 283 | >20 |  |  |  |  |

Values outside the reference range are shown with blue indicating below the reference range and red indicating above the reference range.

Subsequently, the patient was readmitted for infectious diarrhea, oral ulcers, and severe malnutrition. The clinical course was further complicated by chronic diarrhea and recurrent upper respiratory tract infections, with documented pathogens including SARS-CoV-2 and cytomegalovirus (CMV), the latter complicated by CMV retinitis, indicating progressive cellular immunodeficiency.

Recently, the patient developed recurrent severe pneumonia with persistent radiological abnormalities that showed only partial resolution despite antimicrobial therapy. Concurrently, the patient was diagnosed with Salmonella typhimurium enteritis, sinusitis, and developmental delay. During hospitalizations, the patient exhibited recurrent diarrhea, elevated transaminase levels, and eczematous skin lesions, which were managed with antibiotics.

The patient recently underwent allogeneic hematopoietic stem cell transplantation (HSCT) at our center and is currently in the early post-transplant period, with no evidence of graft-versus-host disease (GVHD) to date.

### Identification of compound heterozygous GTF3A mutations as the genetic etiology

Whole-exome sequencing was performed in the patient and both parents to identify the causal gene. The patient carried compound heterozygous mutations in GTF3A: a maternally inherited missense variant c.800T>G (p.I267S) and a paternally inherited frameshift variant c.1085-1088dup (p.L364Yfs*37) **(Fig. 2A-B)**. The I267S variant exhibited a Combined Annotation Dependent Depletion (CADD) score of 25, and the affected isoleucine residue at position 267 is highly conserved across species, suggesting that this missense change is likely deleterious **(Fig. 2C-D)**. The p.L364Yfs*37 variant is a frameshift mutation that alters the reading frame downstream of the duplication and extends the open reading frame, resulting in a delayed termination codon (stop-loss), presumably producing a prolonged protein product with an aberrant C-terminal sequence. These two variants are located in the ZF8 domain and the C-terminal region of GTF3A, respectively **(Fig. 2E)**. Western blot analysis of peripheral blood mononuclear cells demonstrated markedly reduced TFIIIA protein expression in the patient compared to healthy controls, with partially reduced expression in the father and normal expression in the mother **(Fig. 2F)**. GTF3A mutations have recently been reported to cause herpes simplex encephalitis by disrupting biogenesis of the host-derived RIG-I ligand RNA5SP141 (*11*), further supporting the critical role of this gene in human immunity. Collectively, these genetic and protein expression data strongly support that the compound heterozygous GTF3A mutations identified in this patient are pathogenic and account for the disease phenotype.

**Figure 2.**
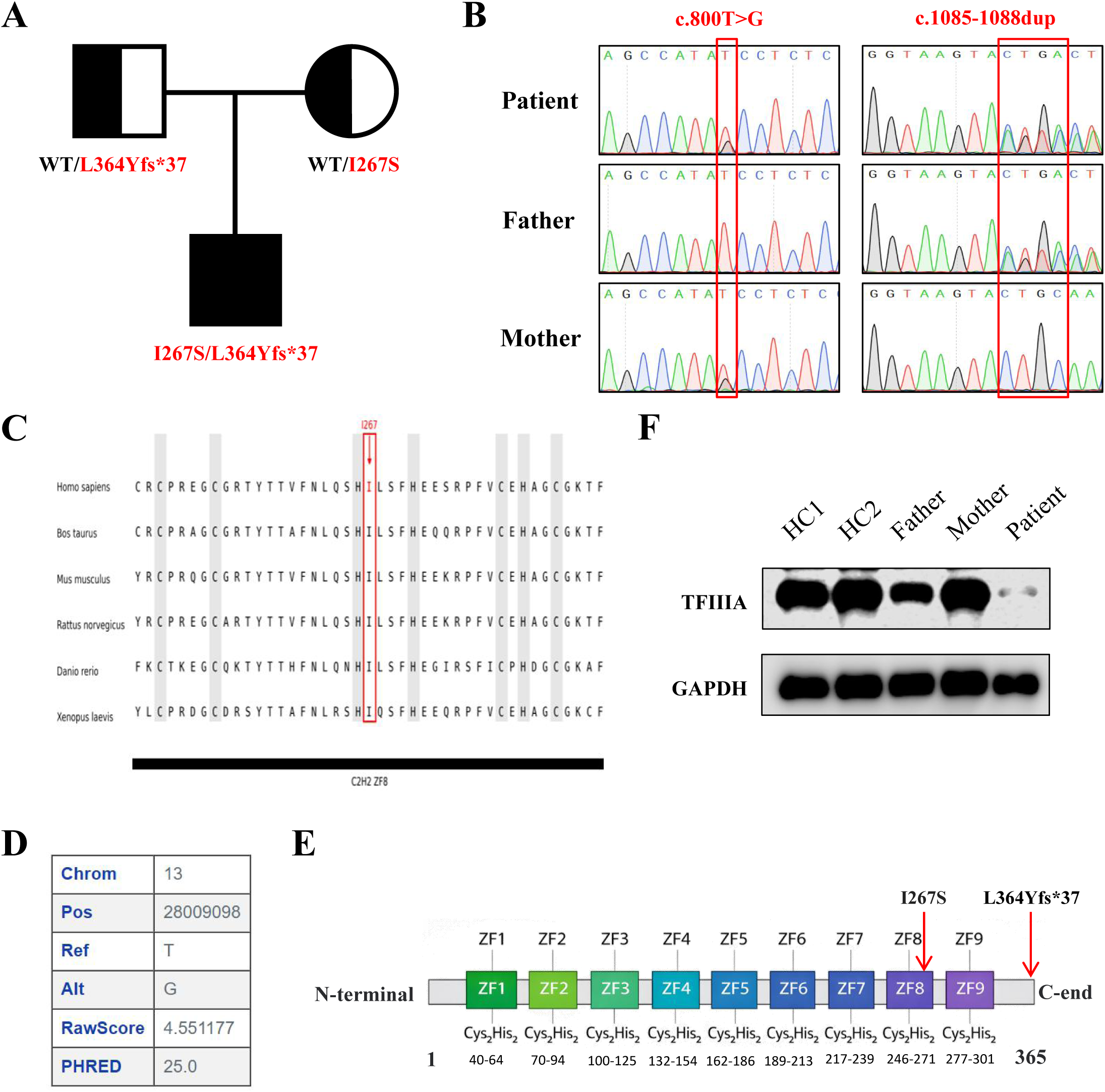
Identification and functional characterization of biallelic GTF3A mutations. (A) Pedigree of the family showing the patient and carrier parents. (B) Sanger sequencing chromatograms confirming the compound heterozygous mutations (c.800T>G, p.I267S and c.1085-1088dup, p.L364Yfs*37) in the patient and respective carrier status in parents. (C) Amino acid conservation analysis of the I267 residue across multiple species. (D) Combined Annotation Dependent Depletion (CADD) scores for the identified variants, with the c.800T>G, p.I267S variant exhibiting a CADD score of 25. (E) Schematic representation of the GTF3A protein domains showing the location of the identified mutations: I267S lies within the ZF8 domain, whereas L364Yfs*37 is located in the C-terminal region and represents a stop-loss mutation that extends the open reading frame. (F) Western blot analysis of TFIIIA protein expression in PBMCs from the patient, parents, and healthy controls.

### Severe T-cell developmental and functional defects

The patient presented with persistent peripheral leukopenia and lymphopenia **(Fig. 1C)**. We next performed a comprehensive phenotypic and functional characterization of T lymphocytes. The absolute counts of both total CD3⁺ T cells and CD19⁺ B cells were below the normal reference ranges **(Table 1)**. Flow cytometric analysis revealed a reduced proportion of CD3⁺ T cells with a concomitant relative increase in CD19⁺ B-cell frequencies. Absolute counts of both CD4⁺ and CD8⁺ T-cell subsets were diminished with inverted CD4/CD8 ratio, and CD56⁺ NK cell counts were also reduced **(Table 1)**. Thymic output was markedly impaired, as evidenced by profoundly reduced TREC levels, whereas KREC levels remained relatively normal. Consistent with this finding, the proportion of CD31⁺ recent thymic emigrants (RTEs) was decreased in both CD4⁺ and CD8⁺ compartments **(Fig. 3A)**. Analysis of peripheral CD4⁺ helper T-cell subsets revealed a reduced frequency of Tfh cells, a relative increase in Tfr cells, and a marked expansion of Treg cells, driven by increases in both CD45RA⁻FOXP3ʰⁱᵍʰ activated Tregs and CD45RA⁻FOXP3ˡᵒʷ non-suppressive Tregs **(Fig. 3B-E)**. In contrast, the proportion of mucosal-associated invariant T (MAIT) cells remained within the normal range **(Fig. 3F)**. Collectively, these findings indicate a profound T-cell developmental defect with impaired thymic output, altered helper T-cell subset distribution, and a regulatory T-cell compartment shift, collectively contributing to the patient’s combined immunodeficiency phenotype.

**Figure 3.**
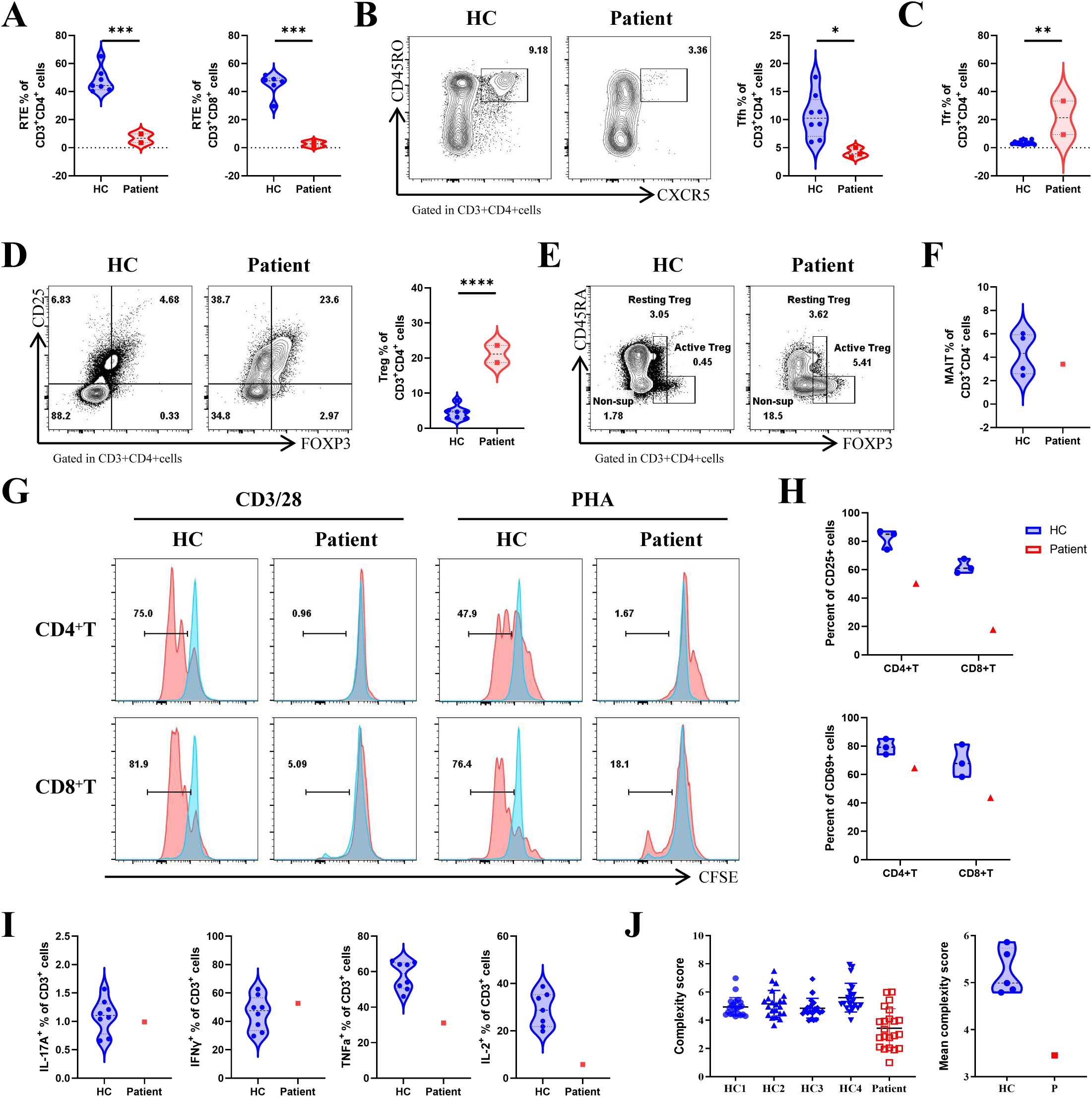
Impaired thymic T cell output, altered helper T cell subsets, and defective T cell function. (A) Flow cytometric analysis of CD31⁺ recent thymic emigrants (RTEs) within CD4⁺ and CD8⁺ compartments, alongside TREC quantification, showing markedly reduced thymic output in the patient compared with healthy controls (HC). (B-E) Flow cytometric frequencies of peripheral CD4⁺ helper T cell subsets, including Tfh (B), Tfr (C), and Treg cells (D), with expanded Tregs driven by increased CD45RA⁻FOXP3ʰⁱᵍʰ activated Tregs and CD45RA⁻FOXP3ˡᵒʷ non-suppressive Tregs (E). Data are representative of two independent experiments. (F) Flow cytometric analysis of mucosal-associated invariant T (MAIT) cell proportions, which remained within the normal range. (G) CFSE-based proliferation assay of patient T cells following anti-CD3/CD28 stimulation, showing profoundly impaired proliferative responses compared with HC. (H) Flow cytometric analysis of surface upregulation of activation markers CD25 and CD69 after TCR stimulation, showing reduced expression in patient T cells. (I) Intracellular cytokine staining for IFN-γ, TNF-α, and IL-2 production upon PMA/ionomycin stimulation, showing preserved apoptosis but markedly diminished cytokine production in patient T cells. (J) TCR Vβ repertoire diversity assessed by spectratyping, showing restricted diversity with monoclonal or oligoclonal peaks in the patient compared with the Gaussian distribution observed in HC. Statistical significance was determined using unpaired two-tailed Student’s t-test or Mann-Whitney U test as appropriate. Error bars represent mean ± SD. HC, healthy control; ns, not significant.

We next assessed the functional competence of patient T cells. Upon stimulation with anti-CD3/CD28 antibodies or phytohemagglutinin (PHA), patient T cells exhibited profoundly impaired proliferative responses compared with healthy controls, as measured by CFSE dilution **(Fig. 3G)**. This proliferative defect was accompanied by reduced surface upregulation of the early activation markers CD25 and CD69 **(Fig. 3H)**, indicating that the T-cell activation threshold was compromised. We next examined whether the T cell lymphopenia also reflected increased susceptibility to cell death. Patient CD4⁺ and CD8⁺ T cells exhibited significantly elevated spontaneous apoptosis compared with healthy controls. Upon Fas engagement, patient T cells showed enhanced apoptosis, and stimulation with PMA and ionomycin—which imposes strong metabolic and signaling demands—also triggered markedly increased cell death **(Supplementary Fig. 1A-B)**. In striking contrast, apoptosis induced by TCR restimulation with anti-CD3/CD28, which engages the classical activation-induced cell death (AICD) pathway, remained comparable between patient and control T cells **(Supplementary Fig. 1)**. Thus, TFIIIA deficiency renders T cells inherently more prone to death through both intrinsic and extrinsic apoptotic pathways, likely reflecting diminished translational reserve, without compromising the specific TCR-driven apoptotic program. Furthermore, cytokine production was markedly impaired: upon PMA/ionomycin stimulation, patient T cells produced significantly reduced amounts of TNF-α and IL-2 compared with healthy controls, as measured by intracellular cytokine staining **(Fig. 3I)**. This functional impairment was further underscored by a severely restricted TCR repertoire: TCR Vβ spectratyping revealed a skewed repertoire dominated by monoclonal or oligoclonal peaks, indicative of profound clonal restriction **(Fig. 3J)**. Collectively, these findings indicate that patient T cells harbor intrinsic defects in activation, proliferation, and cytokine production, with a markedly contracted TCR repertoire and heightened susceptibility to both spontaneous and stress-induced apoptosis. This combined deficit in T cell fitness—encompassing impaired expansion and diminished survival—likely synergizes to drive the progressive T cell depletion and recurrent infections observed in this patient.

### Impaired antibody production and functional defects in patient-derived B cells

Our patient exhibited severe hypogammaglobulinemia with concomitant reduction in absolute B-cell counts. Analysis of peripheral B-cell subsets revealed a significantly decreased frequency of switched memory B cells and a reciprocal increase in naïve B cells compared with age-matched healthy controls **(Fig. 4A)**. The proportions of IgG⁺ and IgA⁺ switched memory B cells, as well as IgM⁺ unswitched memory B cells, were markedly diminished within the total CD19⁺ B-cell compartment **(Fig. 4B)**. Plasmablast frequencies were reduced, while transitional B cells were significantly expanded relative to controls **(Fig. 4C)**. In contrast, the frequency of CD21^low^ B cells remained comparable to that of healthy controls **(Fig. 4D)**. Upon combined stimulation with BCR, CD40L, IL-4, and IL-21, sorted total B cells from patients showed impaired differentiation into plasmablasts (CD27⁺CD38⁺), accompanied by markedly reduced secretion of IgG, IgM, and IgA into culture supernatants **(Fig. 4E-F)**. This functional defect was intrinsic to the B-cell compartment, as similar impairment was recapitulated in differentiation assays using sorted naïve B cells **(Fig. 4G-H)**, which also exhibited profound proliferative defects under the same stimulation conditions **(Fig. 4I)**. Collectively, these findings indicate that patient B cells harbor an intrinsic differentiation defect that results in impaired humoral immune responses.

**Figure 4.**
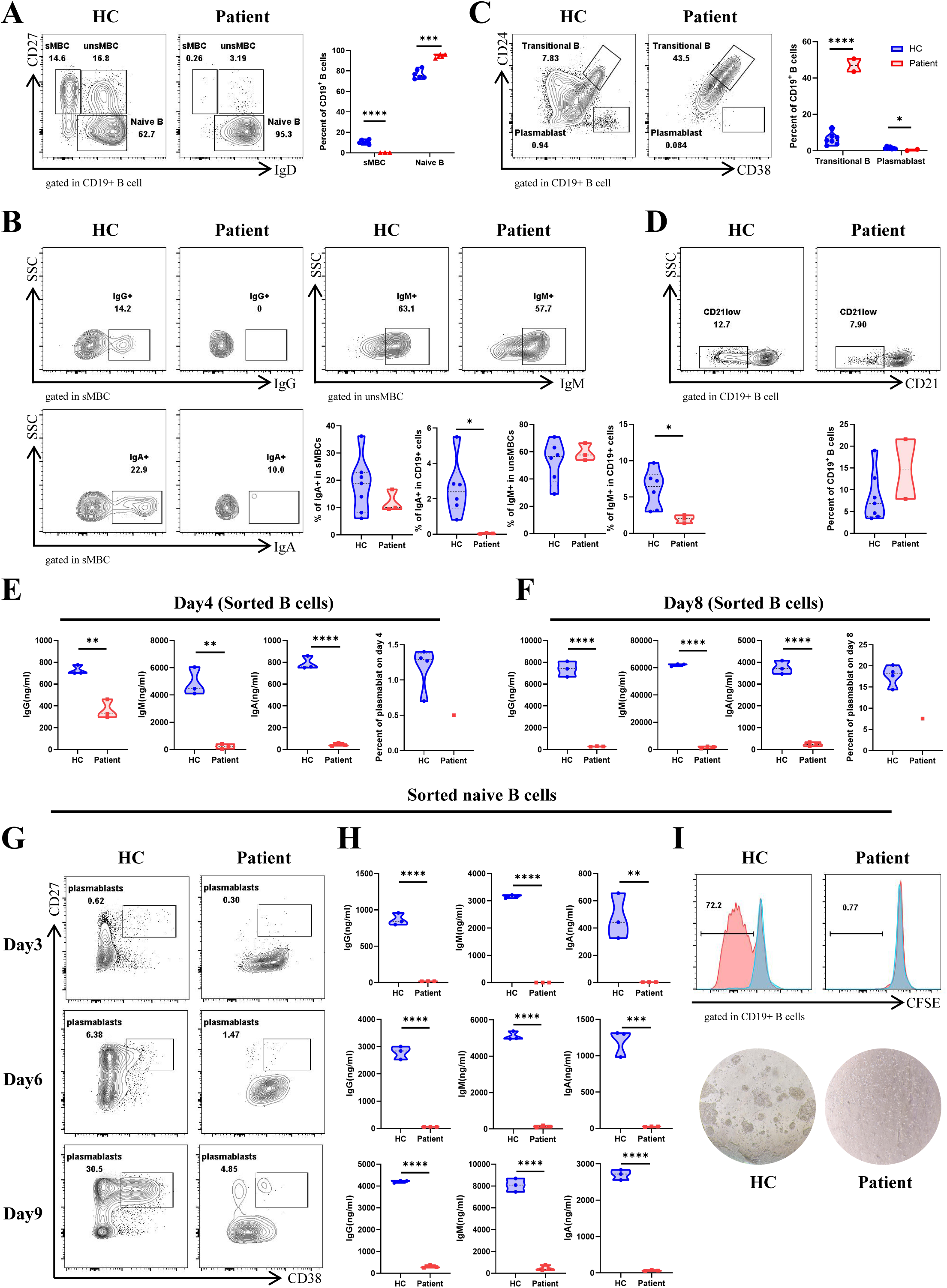
Reduced memory B cells and impaired B cell proliferation and differentiation. (A) Flow cytometric analysis of peripheral B-cell subsets showing decreased switched memory B cells and increased naïve B cells in the patient compared with HC. (B) Proportions of IgG⁺、IgA⁺ switched and IgM⁺ unswitched memory B cells within total CD19⁺ B cells. (C) Flow cytometric analysis of plasmablasts and transitional B cells. (D) Flow cytometric analysis of CD21^low^ B cells in CD19^+^ cells. (E-F) In vitro differentiation of sorted total B cells upon BCR/CD40L/IL-4/IL-21 stimulation, showing impaired plasmablast generation and reduced IgG, IgM, and IgA secretion. (G-H) Differentiation assays using sorted naïve B cells cultured for 3, 6, and 9 days, showing impaired plasmablast differentiation (G) and reduced IgG, IgM, and IgA secretion (H) at all time points, recapitulating the intrinsic differentiation defect. (I) Proliferative responses of sorted naïve B cells under the same stimulation conditions. Data are representative of at least two independent experiments. Error bars represent mean ± SD. HC, healthy control. Statistical significance was determined using unpaired two-tailed Student’s t-test (*p < 0.05, **p < 0.01, ***p < 0.001; ns, not significant).

### GTF3A variants disrupt 5S rRNA and RNA5SP141 pseudogene transcription

To further evaluate the pathogenicity of the GTF3A mutations, we performed a series of in vitro cell-based assays. In overexpression experiments using Flag-tagged constructs in HEK293T cells, the L364Yfs*37 mutant produced a protein band with an increased molecular weight but markedly reduced expression levels, whereas the I267S mutant exhibited expression levels comparable to wild-type (WT) TFIIIA **(Fig. 5A)**. Given the central role of TFIIIA in mediating 5S rRNA transcription, we observed that both mutants exhibited significantly reduced transcription of 5S rRNA and the RNA5SP141 pseudogene **(Fig. 5B)**, the latter of which has recently been identified as a direct target of TFIIIA (*11*). Consistently, impaired transcription of 5S rRNA and RNA5SP141 was also observed in patient-derived PBMCs **(Fig. 5C)**.

**Figure 5.**
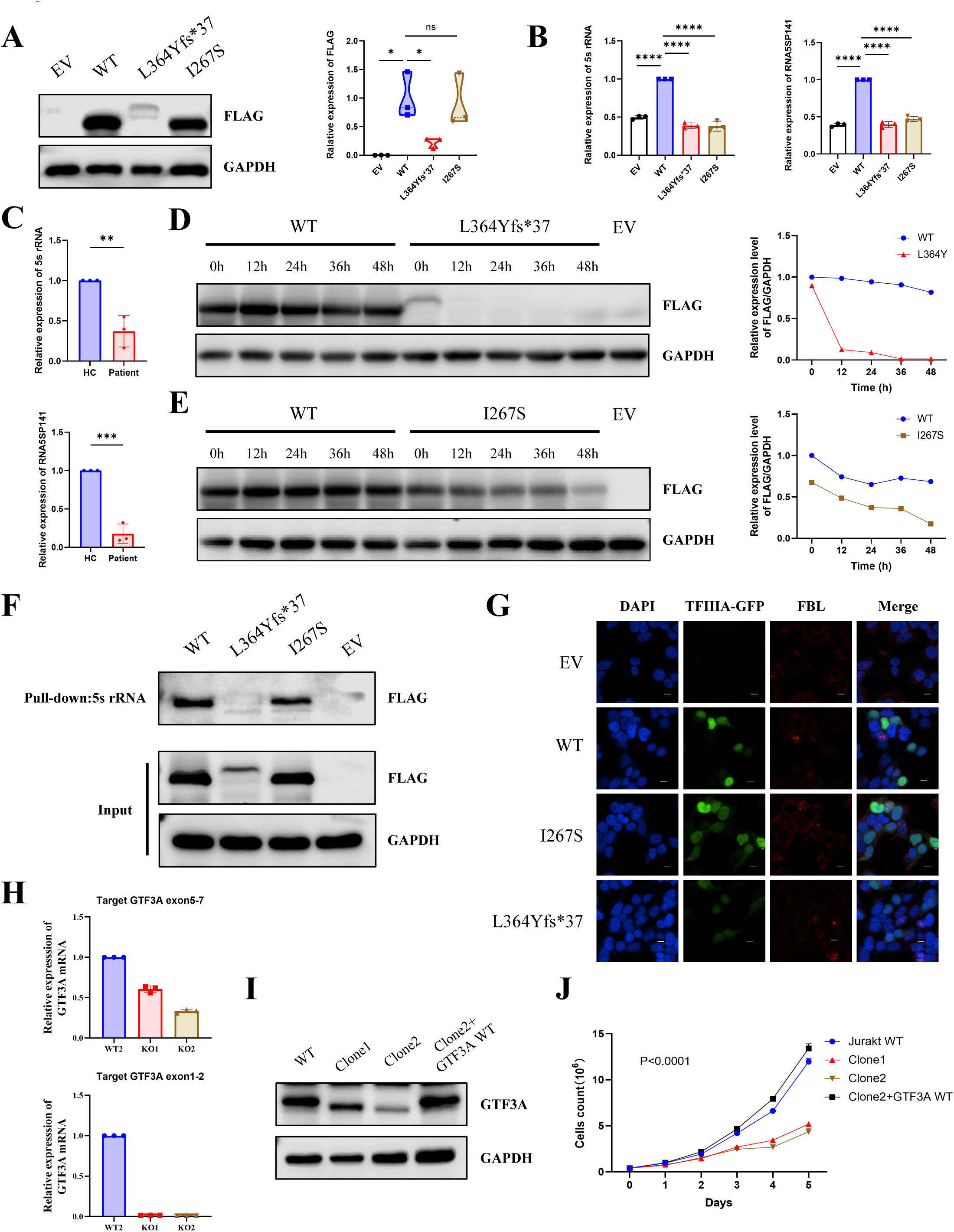
Impaired TFIIIA protein stability, reduced 5S rRNA and RNA5SP141 transcription, and defective proliferation in GTF3A-knockdown Jurkat cells. (A) Western blot analysis of Flag-tagged WT and mutant GTF3A overexpression in HEK293T cells. (B-C) RT-qPCR analysis of 5S rRNA and RNA5SP141 transcript levels in HEK293T cells overexpressing WT or mutant GTF3A and in patient-derived PBMCs. (D-E) Cycloheximide (CHX) chase assay assessing TFIIIA protein stability. (F) RNA pull-down assay using biotinylated 5S rRNA as bait. (G) Confocal microscopy of GFP-tagged WT and mutant TFIIIA in HEK293T cells. (H-I) GTF3A mRNA and TFIIIA protein expression levels in parental Jurkat cells, Clone1 (heterozygous knockout), and Clone2 (further knockdown). (J) Proliferative defects in Clone1 and Clone2 assessed by live-cell counting over serial culture. Lentiviral reconstitution of WT GTF3A in Clone2 restored TFIIIA protein expression and fully rescued the proliferative defect. Data are representative of at least three independent experiments. Error bars represent mean ± SD. Statistical significance was determined using unpaired two-tailed Student’s t-test or one-way ANOVA as appropriate (*p < 0.05, **p < 0.01, ***p < 0.001; ns, not significant).

Given that the L364Yfs*37 variant introduces an aberrant C-terminal extension of 34 amino acids that may lead to protein misfolding and consequent instability, we further quantified the impact of GTF3A mutations on protein stability using a cycloheximide (CHX) chase assay. The L364Yfs*37 mutant exhibited pronounced instability, suggesting that its functional impairment is likely driven by protein instability or conformational disruption **(Fig. 5D)**, whereas the I267S mutation also led to a relatively milder degree of protein instability **(Fig. 5E)**.

Beyond its role in 5S rDNA transcription, TFIIIA functions as a chaperone by directly binding nascent 5S rRNA, thereby promoting its stability and facilitating proper subcellular trafficking toward nucleolar ribosome assembly sites (*12*). To assess whether patient-derived GTF3A variants disrupt this RNA-binding function, we performed an RNA pull-down assay using biotinylated 5S rRNA as bait. The L364Yfs*37 mutant exhibited significantly reduced RNA-binding capacity compared with WT TFIIIA, whereas the I267S mutant showed no appreciable difference **(Fig. 5F)**.

Furthermore, the molecular functions of TFIIIA critically depend on its subcellular localization. TFIIIA is synthesized in the cytoplasm and subsequently imported into the nucleus via defined nuclear localization signal (NLS) regions (*13*). In WT cells expressing GFP-tagged TFIIIA, TFIIIA-GFP was predominantly localized to the nucleus. However, no obvious alterations in nuclear localization were observed for either mutant, although TFIIIA L364Yfs*37-GFP displayed weaker expression, likely due to protein instability and accelerated degradation **(Fig. 5G)**.

### TFIIIA dosage insufficiency impairs Jurkat cell proliferation

To dissect the causal relationship between GTF3A mutations and the clinical T cell immunodeficiency phenotype, we sought to generate GTF3A-deficient Jurkat cells using CRISPR-Cas9-mediated genome editing. Despite extensive single-cell cloning several times, we consistently failed to obtain GTF3A homozygous knockout clones, a finding consistent with the essential role of TFIIIA in 5S rRNA transcription and ribosome assembly, suggesting that complete GTF3A loss may be lethal.

Unexpectedly, we recovered a heterozygous mutant subclone (Clone1). Sanger sequencing revealed a large deletion of 10,809 bp accompanied by a 5-bp insertion on one allele, resulting in a frameshift and complete loss of function, whereas the other allele carried an in-frame deletion **(Supplementary Fig. 2A-C)**. Quantitative PCR confirmed that Clone1 exhibited approximately 50% reduction in GTF3A transcript levels relative to wild-type controls **(Fig. 5H)**, and immunoblotting demonstrated expression of a truncated TFIIIA protein with a lower molecular weight at approximately 50% of wild-type levels **(Fig. 5I)**.

To achieve more profound TFIIIA depletion, we subjected Clone1 to a second round of CRISPR editing. Although homozygous knockout remained unattainable, we successfully isolated an additional subclone (Clone2) with further diminished TFIIIA expression **(Supplementary Fig. 2D-F)**. Compared with Clone1, Clone2 showed an additional ∼50% reduction in both GTF3A mRNA and protein levels, retaining only ∼25% of baseline expression **(Fig. 5H-I)**. Phenotypic analysis revealed that both Clone1 and Clone2 exhibited marked proliferative defects **(Fig. 5J)**. Critically, lentiviral reconstitution of wild-type GTF3A in severe mutated Clone2 restored TFIIIA protein expression and fully rescued the proliferative defect **(Fig. 5I-J)**.

Collectively, these data indicate that TFIIIA plays an essential and nonredundant role in Jurkat cell survival. Complete loss is likely lethal, whereas residual low-level TFIIIA expression, although compatible with basal viability, is insufficient to support normal cell proliferation. These findings precisely recapitulate the proliferative failure of patient-derived lymphocytes ex vivo.

## Discussion

The identification of GTF3A as a monogenic cause of combined immunodeficiency expands the growing family of ribosomopathies that manifest as primary immunodeficiencies and establishes TFIIIA as an essential determinant of human lymphocyte fitness. Our findings demonstrate that biallelic GTF3A mutations in a CID patient disrupt 5S rRNA transcription, impair TFIIIA protein stability and RNA-binding capacity, and culminate in intrinsic proliferative defects in both T and B lymphocytes. Through sequential CRISPR engineering in Jurkat cells, we further show that TFIIIA is required for cell survival in a dose-dependent manner and that complete loss is incompatible with viability. Moreover, patient T cells exhibited heightened susceptibility to both spontaneous and stress-induced apoptosis, indicating that TFIIIA insufficiency compromises lymphocyte fitness not only by limiting proliferative expansion but also by impairing basal survival. Collectively, these data position TFIIIA deficiency within the ribosomopathy spectrum and uncover a nonredundant role for 5S rRNA biogenesis in adaptive immunity.

The conceptual link between ribosome dysfunction and primary immunodeficiency has been established through several prototypical ribosomopathies. Cartilage-hair hypoplasia, caused by mutations in RMRP encoding the RNA component of the mitochondrial RNA-processing endonuclease, impairs pre-ribosomal RNA processing and manifests with T cell lymphopenia and variable humoral defects (*14,15*). Shwachman-Diamond syndrome, resulting from SBDS mutations, is likewise characterized by bone marrow failure and immune dysfunction (*16*). Diamond-Blackfan anemia, associated with mutations in ribosomal protein genes such as RPL5 and RPL11, presents with erythroid aplasia but can also affect lymphocyte compartments (*17,18*). Our study adds GTF3A to this list, but with a critical distinction: whereas most ribosomopathies affect broadly expressed ribosomal components and produce pleiotropic phenotypes, TFIIIA is uniquely dedicated to 5S rRNA synthesis. This functional specialization may explain why GTF3A deficiency produces a relatively restricted immunologic phenotype compared with the multisystem involvement seen in other ribosomopathies; nevertheless, the severe T cell lymphopenia and humoral failure in our patient underscore the particular vulnerability of the lymphoid lineage to 5S rRNA insufficiency.

The lymphocyte-intrinsic nature of the defects we observed is supported by multiple lines of evidence. Patient T cells exhibited profoundly impaired proliferative responses upon TCR stimulation, accompanied by diminished activation marker upregulation and severely contracted TCR repertoire diversity. Patient B cells, both total and naïve subsets, showed impaired plasmablast differentiation and reduced immunoglobulin secretion in vitro. Furthermore, patient T cells displayed increased spontaneous apoptosis and enhanced susceptibility to Fas-mediated and stress-induced cell death, whereas TCR-restimulation-induced AICD remained intact—indicating that the increased death vulnerability reflects a global reduction in the apoptotic threshold rather than a specific defect in TCR-driven death programs. These defects were recapitulated in CRISPR-engineered Jurkat cells, where graded TFIIIA depletion impaired proliferation in a dose-dependent manner and wild-type GTF3A reconstitution fully rescued the phenotype. The inability to obtain GTF3A homozygous knockout clones—despite extensive single-cell cloning—further indicates that TFIIIA is essential for Jurkat cell survival. Collectively, these observations establish that TFIIIA deficiency acts cell-intrinsically to disrupt lymphocyte homeostasis, encompassing both proliferative failure and enhanced apoptosis, rather than through extrinsic microenvironmental factors.

The mechanistic basis of TFIIIA-dependent lymphocyte fitness is rooted in its dual function as both a transcriptional activator and an RNA chaperone. TFIIIA initiates RNA polymerase III–dependent transcription at the 5S rDNA internal control region and simultaneously binds nascent 5S rRNA transcripts, chaperoning them toward proper incorporation into the 60S large ribosomal subunit (*19–20*). The patient-derived I267S and L364Yfs*37 variants impaired both of these activities: 5S rRNA and RNA5SP141 transcription were reduced in patient PBMCs and in overexpression assays; RNA pull-down revealed diminished 5S rRNA binding capacity for the L364Yfs37 mutant, whereas the I267S mutant retained binding comparable to wild-type; and cycloheximide chase demonstrated accelerated protein degradation, particularly for the L364Yfs*37 frameshift mutant. The L364Yfs*37 variant, which introduces a 34-amino acid aberrant C-terminal extension, likely disrupts the C-terminal domain critical for transcription activation, whereas the I267S missense change within the ZF8 domain may compromise DNA-binding specificity. The resulting reduction in 5S rRNA availability would be expected to impair ribosome assembly and translational capacity, which may be particularly detrimental to lymphocytes given their requirement for rapid proliferation upon antigen encounter.

An intriguing dimension of TFIIIA biology relates to its transcriptional control of the 5S ribosomal RNA pseudogene 141 (RNA5SP141), which was recently identified as an endogenous RIG-I ligand that contributes to antiviral innate immune responses (*11*). GTF3A mutations have been shown to impair RNA5SP141 transcription and abrogate RIG-I activation upon HSV-1 infection (*11*). Our patient, who suffered from recurrent infections including CMV pneumonia with Pneumocystis jirovecii co-infection and SARS-CoV-2, did not present with HSV-1 encephalitis, suggesting that the immunological consequences of GTF3A mutations may extend beyond HSV-1 susceptibility to a broader CID phenotype. This raises the possibility that TFIIIA deficiency compromises immunity through at least two distinct mechanisms: a cell-intrinsic proliferative defect in lymphocytes driven by impaired ribosome biogenesis, and a cell-intrinsic innate immune defect in antiviral sensing mediated by RNA5SP141 downregulation. The relative contribution of each mechanism to the clinical phenotype likely depends on the specific mutations, residual TFIIIA activity, and environmental infectious pressures.

The severe proliferative defects observed in both T and B cells from the patient may represent the key reason why the adaptive immune system is particularly sensitive to TFIIIA insufficiency. Upon antigen stimulation, T and B cells must rapidly exit quiescence and enter the cell cycle, initiating massive protein synthesis to support clonal expansion and effector differentiation. This process places extraordinary demands on ribosomal functional reserve (*21*). TFIIIA-deficient lymphocytes, facing insufficient 5S rRNA supply, may fail to meet the demands of ribosome assembly and translation when challenged with antigen, thereby precluding effective proliferation. Consistent with this notion, patient T cells showed profoundly impaired proliferation upon anti-CD3/CD28 or PHA stimulation, accompanied by reduced upregulation of CD25 and CD69. Patient B cells similarly exhibited proliferative failure and impaired plasmablast differentiation upon combined BCR/CD40L/IL-4/IL-21 stimulation. Of note, even at baseline, the peripheral T cell TCR Vβ repertoire in the patient displayed marked oligoclonal restriction, likely reflecting compensatory peripheral homeostatic proliferation of residual T cells after severe thymic output failure; however, these compensatory cells remained functionally compromised due to TFIIIA insufficiency and ultimately failed to protect against infections.

The increased apoptotic susceptibility of patient T cells provides a complementary explanation for the profound T cell lymphopenia. The enhanced spontaneous apoptosis, together with heightened sensitivity to Fas engagement and PMA/ionomycin-induced stress, points to a globally lowered apoptotic threshold. This phenotype is consistent with the notion that reduced translational capacity—resulting from 5S rRNA insufficiency—compromises the synthesis of short-lived anti-apoptotic proteins, such as MCL-1 and BCL-2, which are critical for maintaining lymphocyte survival (*22,23*). Conversely, the normal AICD response upon TCR restimulation indicates that the proximal TCR signaling machinery and the FAS/FASL execution pathway remain functionally intact. Thus, TFIIIA deficiency does not broadly disrupt all death programs but instead renders T cells inherently more fragile under basal conditions and in response to extrinsic death signals, while preserving the specific activation-induced death pathway that is essential for peripheral immune tolerance. This heightened basal vulnerability, together with the profound proliferative defect, likely synergizes to drive the severe T cell depletion observed in the patient.

The B cell phenotype in our patient adds another layer to the understanding of TFIIIA function in humoral immunity. The patient exhibited pan-hypogammaglobulinemia with markedly reduced IgG, IgM, and IgA. Peripheral B cell subset analysis revealed a significant reduction in switched memory B cells and plasmablasts, with reciprocal expansion of naïve and transitional B cells. Patient B cells showed impaired differentiation into plasmablasts and reduced immunoglobulin secretion upon stimulation, defects that were recapitulated in sorted naïve B cells. These findings indicate that TFIIIA deficiency disrupts B cell maturation at the germinal center transition and terminal differentiation stages. B cells undergo somatic hypermutation and class-switch recombination in germinal centers, followed by differentiation into plasma cells, a process that requires extensive cell division and protein synthesis. Given that plasma cells are the main producers of antibodies, their differentiation process is particularly reliant on ribosomal function (*24*), making interference by TFIIIA deficiency expected. The expansion of transitional B cells may reflect a bottleneck at the maturation checkpoint in the bone marrow, whereas the depletion of switched memory compartments points to impaired T-dependent humoral responses, likely compounded by the concurrent Tfh deficiency.

Collectively, our study establishes GTF3A as a novel CID-causing gene and positions TFIIIA deficiency within the expanding spectrum of ribosomopathies that manifest as primary immunodeficiencies. The lymphocyte-intrinsic proliferative defects, enhanced apoptotic susceptibility, and humoral failure observed in our patient jointly underscore the nonredundant role of 5S rRNA biogenesis in adaptive immunity. These findings broaden the genetic landscape of inborn errors of immunity and suggest that GTF3A should be considered in the molecular diagnosis of patients with unexplained CID, particularly those presenting with a T-B⁺NK⁺ immunophenotype and recurrent infections.

## Materials and methods

### Genetic Diagnosis

Peripheral blood and DNA samples from the patient were sent to MyGenostics (Beijing, China) for Whole-exome sequencing (WES) (*25*). Mutations in the *GTF3A* gene were confirmed by Sanger sequencing. The primers for the PCR as showed in supplementary materials.

### Statistics

Statistical analysis was performed using GraphPad Prism 8.0 (GraphPad Software, La Jolla, CA, USA). Flow cytometry data were analyzed with FlowJo 10.3 (TreeStar Inc., Ashland, OR, USA). The specific statistical tests applied for each experiment are indicated in the corresponding figure legends. A P value < 0.05 was considered statistically significant.

**Additional methods are available in the Supplementary Materials.**

## Author Contributions

YF.A., and XD.Z. supervised this study, reviewed and revised the manuscript. L.Y., HL.L., QL.W., and JJ.W. contributed equally to this work as co-first authors. YL.L., LW.Z.,

WH.L., LN.Z., YJ.J., Y.D., and Q.Z. performed some experiments and essential help for clinical management and follow-up of the patient. All authors contributed to the final version of the manuscript and approved submission of the final version.

## Acknowledgments

We would like to thank the patient, the unaffected controls and their parents for their participation.

## Ethics statement

The study was performed following the Declaration of Helsinki and approved by the ethics committee of Children’s Hospital of Chongqing Medical University (Chongqing, China). Written informed consent for participation in this study were obtained from patient’s parents.

## Data availability

The dataset described in this study is available from the corresponding authors upon reasonable request.

## Reference

1. Notarangelo LD, Bacchetta R, Casanova JL, Su HC. Human inborn errors of immunity: An expanding universe. Sci Immunol. 2020;5(49):eabb1662. doi:10.1126/sciimmunol.abb1662

2. Poli MC, Aksentijevich I, Bousfiha AA, et al. Human inborn errors of immunity: 2024 update on the classification from the International Union of Immunological Societies Expert Committee. J Hum Immun. 2025;1(1):e20250003. Published 2025 Apr 15. doi:10.70962/jhi.20250003

3. Meyts I, Bosch B, Bolze A, et al. Exome and genome sequencing for inborn errors of immunity. J Allergy Clin Immunol. 2016;138(4):957–969. doi:10.1016/j.jaci.2016.08.003

4. Bousfiha AA, Jeddane L, Moundir A, et al. The 2024 update of IUIS phenotypic classification of human inborn errors of immunity. J Hum Immun. 2025;1(1):e20250002. Published 2025 Apr 15. doi:10.70962/jhi.20250002

5. Sulima SO, Hofman IJF, De Keersmaecker K, Dinman JD. How Ribosomes Translate Cancer. Cancer Discov. 2017;7(10):1069–1087. doi:10.1158/2159-8290.CD-17-0550

6. Farley-Barnes KI, Ogawa LM, Baserga SJ. Ribosomopathies: Old Concepts, New Controversies. Trends Genet. 2019;35(10):754–767. doi:10.1016/j.tig.2019.07.004

7. Engelke DR, Ng SY, Shastry BS, Roeder RG. Specific interaction of a purified transcription factor with an internal control region of 5S RNA genes. Cell. 1980;19(3):717–728. doi:10.1016/s0092-8674(80)80048-1

8. Pelham HR, Brown DD. A specific transcription factor that can bind either the 5S RNA gene or 5S RNA. Proc Natl Acad Sci U S A. 1980;77(7):4170–4174. doi:10.1073/pnas.77.7.4170

9. Setzer DR, Menezes SR, Del Rio S, Hung VS, Subramanyan G. TFIIIA: a multifunctional zinc finger protein. Gene Expr. 1996;6(2):63–75.

10. Layat E, Probst AV, Tourmente S. Structure, function and regulation of Transcription Factor IIIA: From Xenopus to Arabidopsis. Biochim Biophys Acta. 2013;1829(3-4):274–282. doi:10.1016/j.bbagrm.2012.10.013

11. Naesens L, Muppala S, Acharya D, et al. GTF3A mutations predispose to herpes simplex encephalitis by disrupting biogenesis of the host-derived RIG-I ligand RNA5SP141. Sci Immunol. 2022;7(77):eabq4531. doi:10.1126/sciimmunol.abq4531

12. Lee BM, Xu J, Clarkson BK, et al. Induced fit and “lock and key” recognition of 5S RNA by zinc fingers of transcription factor IIIA. J Mol Biol. 2006;357(1):275–291. doi:10.1016/j.jmb.2005.12.010

13. Wischnewski J, Rudt F, Pieler T. Signals and receptors for the nuclear transport of TFIIIA in Xenopus oocytes. Eur J Cell Biol. 2004;83(2):55–66. doi:10.1078/0171-9335-00358

14. Vakkilainen S. Cartilage-hair hypoplasia: A comprehensive review. J Hum Immun. 2025;1(4):e20250142. Published 2025 Oct 1. doi:10.70962/jhi.20250142

15. Pello E, Kainulainen L, Vakkilainen M, et al. Shorter birth length and decreased T-cell production and function predict severe infections in children with non-severe combined immunodeficiency cartilage-hair hypoplasia. J Allergy Clin Immunol Glob. 2023;3(1):100190. Published 2023 Nov 22. doi:10.1016/j.jacig.2023.100190

16. Gloude NJ, Brundige K, Loveless S, et al. Beyond neutropenia: Immunological evaluation of patients with Shwachman-Diamond syndrome. Br J Haematol. 2026;209(1):180–188. doi:10.1111/bjh.70553

17. Nelson A, Myers K. Shwachman-Diamond Syndrome. In: Adam MP, Bick S, Mirzaa GM, Pagon RA, Wallace SE, Amemiya A, eds. GeneReviews®. Seattle (WA): University of Washington, Seattle; July 17, 2008.

18. Fellmann F, Saunders C, O’Donohue MF, et al. An atypical form of 60S ribosomal subunit in Diamond-Blackfan anemia linked to RPL17 variants. JCI Insight. 2024;9(17):e172475. Published 2024 Aug 1. doi:10.1172/jci.insight.172475

19. Layat E, Probst AV, Tourmente S. Structure, function and regulation of Transcription Factor IIIA: From Xenopus to Arabidopsis. Biochim Biophys Acta. 2013;1829(3-4):274–282. doi:10.1016/j.bbagrm.2012.10.013

20. Tan C, Li W, Wang W. Localized frustration and binding-induced conformational change in recognition of 5S RNA by TFIIIA zinc finger. J Phys Chem B. 2013;117(50):15917–15925. doi:10.1021/jp4052165

21. Sinclair LV, Cantrell DA. Protein Synthesis and Metabolism in T Cells. Annu Rev Immunol. 2025;43(1):343–366. doi:10.1146/annurev-immunol-082323-035253

22. Bose P, Grant S. Mcl-1 as a Therapeutic Target in Acute Myelogenous Leukemia (AML). Leuk Res Rep. 2013;2(1):12–14. doi:10.1016/j.lrr.2012.11.006

23. Youle RJ, Strasser A. The BCL-2 protein family: opposing activities that mediate cell death. Nat Rev Mol Cell Biol. 2008;9(1):47–59. doi:10.1038/nrm2308

24. He X, Zhao J, Adilijiang A, et al. Dhx33 promotes B-cell growth and proliferation by controlling activation-induced rRNA upregulation. Cell Mol Immunol. 2023;20(3):277–291. doi:10.1038/s41423-022-00972-0

25. Yu L, Zhou B, Liu W, et al. Autosomal dominant gain-of-function mutations in LCP1 cause a syndromic neutropenia and immunodeficiency. Genes Dis. 2026;102232. doi:10.1016/j.gendis.2026.102232.

