## Supplemental Figures for "Biallelic GTF3A mutations underline a novel human combined immunodeficiency"

### Supplementary Methods

#### Antibodies and reagents

For immunoblotting, the following antibodies were used: anti-FLAG (Sigma-Aldrich), anti-α-Tubulin (Proteintech), anti-TFIIIA antibody (Proteintech), goat anti-mouse IgG HRP (Zhongshan Golden Bridge), and goat anti-rabbit IgG HRP (Zhongshan Golden Bridge). For flow cytometry, the following anti-human clones were used: CD3-BV421 (HIT3a, BioLegend), CD4-PE-Cy7 (RPA-T4, BioLegend), CD8-PE (BioLegend), CD19-APC (BioLegend), CD45RA-FITC (HI100, BD), CD45RO-APC (UCHL1, BD), CXCR5-BV421 (J252D4, BioLegend), PD-1-FITC (EH12.2H7, BioLegend), ICOS-PE (DX29, BD), CD127-PE (A019D5, BioLegend), CD25-APC (BC96, BioLegend), CXCR3-APC (1C6, BD), CCR6-PE (G034E3, BioLegend), CD56-FITC (BioLegend), CD86-PE (BioLegend), anti-human IgG-FITC (BioLegend), and IgA-PE (Miltenyi Biotec). Fixation/permeabilization kit (Thermo Fisher Scientific) and CellTrace CFSE Cell Proliferation Kit (Invitrogen) were used. Additional reagents included Dual Luciferase Reporter Assay System (Promega), Poly-L-lysine-coated slides (Solaibio), LPS (Sigma), PMA (AbMole), ammonium thiocyanate (Aladdin), CpG ODN (InvivoGen), F(ab')2 goat anti-human IgM (Jackson ImmunoResearch), Ultra-LEAF purified anti-human CD28 and anti-human CD3 (BioLegend), and fetal bovine serum (ExCellBio).

#### Plasmid construction

HEK293T cells were cultured in DMEM supplemented with 10% FBS, 100 U/mL penicillin, and 100 μg/mL streptomycin at 37°C in 5% CO2. Plasmids encoding 3×Flag-tagged wild-type human GTF3A and its mutant variants were constructed. Site-directed mutagenesis was performed using overlap-extension PCR with the wild-type construct as template; all mutations were verified by Sanger sequencing. Transfections were carried out using polyethylenimine (PEI), and cells were harvested 24 h post-transfection for downstream assays.

#### Immunoblotting

Protein lysates were prepared from peripheral blood mononuclear cells (PBMCs) of healthy controls and patients, as well as from HEK293T cells transfected with wild-type or mutant GTF3A plasmids, using lysis buffer supplemented with protease and phosphatase inhibitors. Proteins were resolved by SDS-PAGE, transferred to membranes, blocked, and probed with primary antibodies followed by HRP-conjugated secondary antibodies. Chemiluminescent detection was employed to assess TFIIIA expression levels and, where applicable, phosphorylation status of signaling molecules.

**Cell proliferation assay**

PBMCs from patients and healthy controls were washed with PBS and resuspended at $1 \times 10^6$ cells/mL. Cells were labeled with 5 μM CFSE at 37°C for 10 min in the dark, and the reaction was quenched with cold RPMI 1640 containing 10% FBS, followed by two washes. Labeled cells were cultured under four conditions: unstimulated, PHA stimulation (for T cells), anti-CD3/CD28 stimulation (for T cells), or CpG plus anti-human IgM F(ab')2 (for B cells). After 72 h of culture at 37°C in 5% CO2, cells were stained with CD4-PB, CD8a-PE, CD19-APC, and viability dye L/D780, then analyzed by flow cytometry. Proliferation was assessed by CFSE dilution in each lymphocyte subset.

**Th1, Th2, and Th17 subset analysis**

PBMCs from patients and controls were stained with CD3-Percp, CD4-PE-Cy7, CD45RA-FITC, CXCR5-BV421, PD-1-FITC, ICOS-PE, CD127-PE, CD25-APC, CXCR3-APC, and CCR6-PE. After red blood cell lysis and washing, samples were acquired on a flow cytometer. Frequencies of Th1, Th2, Th17, Tfh, and Tfr cells were determined based on surface marker expression profiles.

**PBMC activation and cytokine measurement**

For T-cell activation marker analysis, PBMCs were stimulated with anti-CD3/CD28 for 48 h, then stained with fluorochrome-conjugated antibodies against CD3, CD4, CD8, CD25, and CD69. The percentages of CD25+ or CD69+ cells within CD4+ or CD8+ subsets were determined by flow cytometry and analyzed with FlowJo.

Intracellular cytokine staining was used to assess IL-17A, IFN-γ, TFN-α, and IL-2 production. For activation, PBMCs were stimulated with PMA (50 ng/mL) and ionomycin (1 μg/mL) for 5 h in the presence of brefeldin A. For intracellular staining, cells were fixed, permeabilized, and stained with fluorochrome-conjugated anti-cytokine antibodies before flow cytometric analysis.

**Plasmablast differentiation assay**

Cryopreserved PBMCs were thawed, and B cells were enriched using a Pan B Cell Isolation Kit or Naive B cells Isolation Kit (STEMCELL). Purified B cells (5 × 10^4 per well) were cultured in RPMI 1640 with 10% FBS, supplemented with IL-2 (100 IU/mL), IL-4 (100 ng/mL), IL-21 (100 ng/mL), IL-10 (25 ng/mL), CD40L (200 ng/mL), and anti-IgM (5 μg/mL). Medium was refreshed every 3 days. Differentiation into plasmablasts was assessed by flow cytometry, and immunoglobulin secretion (IgG, IgM, IgA) in culture supernatants was measured by ELISA (4A Biotech) on days 3, 6, and 9.

**Primers for PCR or qPCR**

**Supplementary table 1. PCR primer sequences for the GTF3A gene**

| Target gene | Primer sequences |
|---|---|
| *GTF3A* c.800T>G DNA F | ATGTACAACAGCATGCCTAGTGAG |
| *GTF3A* c.800T>G DNA R | GCCGGAAAACATTTAAACGCAAAG |
| *GTF3A* c.1088dup DNA F | CGTGAAAAACGGAGTTTGGCCTC |
| *GTF3A* c.1088dup DNA R | TCACTTAACCGACAGCAGCAAT |

**Supplementary table 1. qPCR primer sequences for this study**

| Target gene | Primer sequences |
|---|---|
| *GTF3A* exon1-2 qPCR F | CGGTGTCGTCCTTGACCATC |
| *GTF3A* exon1-2 qPCR R | TTGGCTGCACAAACAAACGG |
| *GTF3A* exon5-7 qPCR F | CCACGAGGGCTATGTATGTCA |
| *GTF3A* exon5-7 qPCR R | CCTTTCTGGGGCATGAGTTTTC |
| *5s rRNA* qPCR F | GCCATACCACCCTGAACG |
| *5s rRNA* qPCR R | GGTATTCCCAGGCGGTCT |
| *RNA5SP141* qPCR F | CTACGGCCATACCACCCTGAAC |
| *RNA5SP141* qPCR R | ATCCAAGTACTAACCAGGCCCAAC |

**Immunofluorescence microscopy**

HEK293T cells grown on glass coverslips in 24-well plates were transfected with GFP-tagged wild-type or mutant GTF3A plasmids. At 24 h post-transfection, cells were fixed with 4% paraformaldehyde for 30 min, permeabilized with 0.1% Triton X-100 for 15 min, and blocked with 5% BSA in PBS. Samples were then incubated overnight at 4°C with anti-fibrillarin primary antibody, followed by fluorophore-conjugated secondary antibody for 1 h at room temperature in the dark. Nuclei were counterstained with DAPI for 15 min, and coverslips were mounted with anti-fade medium. Confocal images were acquired using a Nikon A1 laser scanning microscope and analyzed with NIS-Elements software.

**Apoptosis assay**

T-cell apoptosis was evaluated under spontaneous and induced conditions. PBMCs were cultured in complete RPMI 1640 with 10% FBS for 24–72 h without stimulation (spontaneous) or with plate-bound anti-CD3 (5 μg/mL)/soluble anti-CD28 (5 μg/mL) or FAS ligand (100 ng/mL) for 24–72 h (induced). Apoptotic cells were detected by Annexin V and 7-AAD staining (BD Biosciences) according to the manufacturer's protocol. Cells were acquired by flow cytometry, and early (Annexin V+7-AAD-) and late (Annexin V+7-AAD+) apoptotic populations were quantified using FlowJo software.

All experiments were performed with at least three independent biological replicates, and data are presented as mean ± SD. Statistical significance was assessed using unpaired Student's t-test or one-way ANOVA as appropriate.

**Construction of GTF3A- deficient Jurkat cell lines**

CRISPR-Cas9 mediated ablation of the GTF3A gene was achieved with CRISPR-Cas9 RNP (provided by Haixing Bioscience) containing expression cassettes for hSpCas9 and chimeric guide RNA. To target exon 1~9 of the GTF3A gene, two guide RNA sequence of GAACGCGTCGGCGATGGTCA AGG and ACAGTTGCAGTACTTACCCT TGG were selected through the http://crispr.mit.edu website. RNP complexes was electrotransfected into cells using Neon transfection system according to the manufacturer’s instructions (ThermoFisher Scientific). After two days, single colonies were transferred into 96-well plates. To determine the presence of insertions or deletions (indels) in GTF3A targeted clones, genomic DNA was isolated using a Quick-DNA Miniprep kit (Zymo Research) and PCR amplification was achieved using 2×Taq Master Mix (Dye Plus; Vazyme, P112) of primers flanking exon; Forward: 5’-CTTGCAGCAGAAGCACGTAA-3’; Reverse: 5’-CTTAACCGACAGCAGCAATTCA-3’. After the first round of electroporation, no monoclonal cells harboring biallelic knockout were successfully obtained. Consequently, heterozygous cells were selected for the second round of targeted editing. To target exon 2~6 of the GTF3A gene, two guide RNA sequence of ATACTACCAAGGCGTCCAGG AGG and AGCATGCTAATTGTGCATGG TGG were selected through the http://crispr.mit.edu website. The identification primers for the second round of targeting are as follows: Forward: 5’-GGGCTGCAGTCCATACCTTT-3’; Reverse: 5’-CCCTTTCTGGGGCATGAGTT-3’. PCR products were subjected to Sanger sequencing (GENEWIZ, China). Clones with mutations in both alleles were selected for downstream studies. Cell lines generated using the above strategy includeWT, clone1, and clone2. All clones were maintained under the same conditions as parental cells.

The PCR primer sequences used to obtain Clone 1 from the first round of electroporation are as follows:

| Location | Primer name | Primer sequence (5'->3') |
|---|---|---|
| **Primers for Region 1** | Region 1-F | CTTGCAGCAGAAGCACGTAA |
| | Region 1-R | TGGATGTCAAAGCTGAACGC |
| **Primers for Region 2** | Region 2-F | AAGTCTCACTAGGCATGCTGTT |
| | Region 2-R | CTTAACCGACAGCAGCAATTCA |
| **Primers for Region 3（KO）** | Region 3-F | CTTGCAGCAGAAGCACGTAA |
| | Region 3-R | CTTAACCGACAGCAGCAATTCA |

The PCR primer sequences used to obtain Clone 2 from the second round of electroporation are as follows:

| Location | Primer name | Primer sequence (5'->3') |
|---|---|---|
| **Primers for Region 4** | Region 4-F | GGGCTGCAGTCCATACCTTT |
| | Region 4-R | TTGTCTGCATGGATCCCCAC |
| **Primers for Region 5** | Region 5-F | TAAAGGGAGGCACCTTGCTG |
| | Region 5-R | CCCTTTCTGGGGCATGAGTT |
| **Primers for Region 6（KO）** | Region 6-F | GGGCTGCAGTCCATACCTTT |
| | Region 6-R | CCCTTTCTGGGGCATGAGTT |

### Supplementary Figures

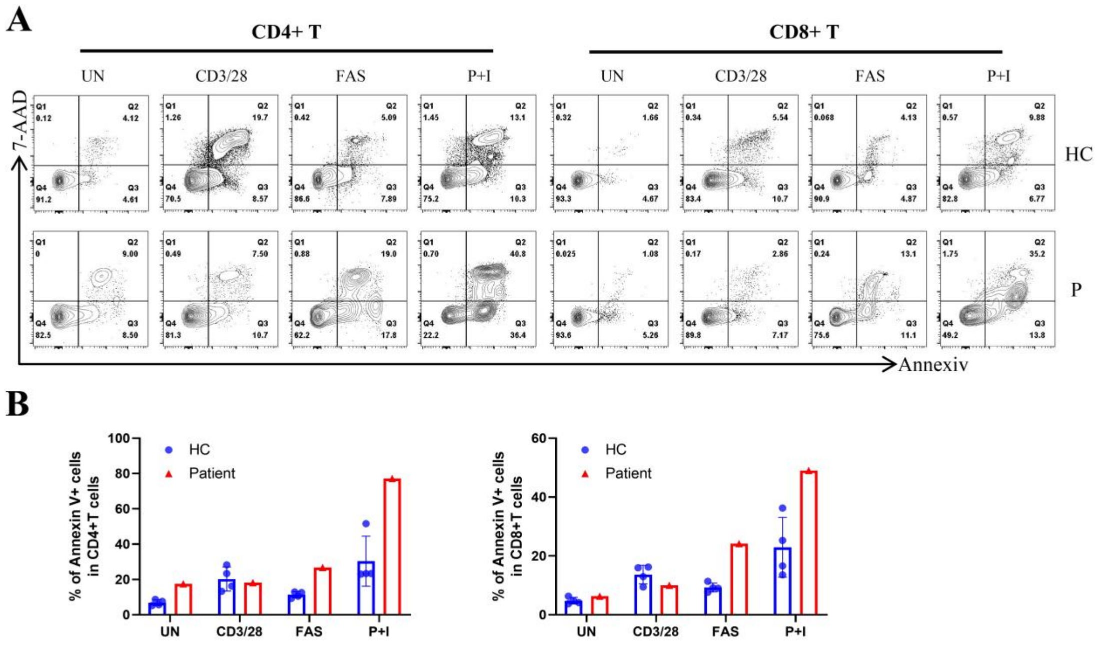


**Supplemental Figure 1. T-cell apoptosis analysis in patients with TFIIIA deficiency.** (A) Representative flow cytometry plots showing Annexin V and 7-AAD staining in $CD4^+$ and $CD8^+$ T cells from a HC and patient P1 and P2 under unstimulated conditions, or following stimulation with anti-CD3/CD28 antibodies or FAS-L or PMA+Ionomycin (P+I) for 72 hours. (B) Summary frequencies of apoptotic cells (Annexin $V^+$) within $CD4^+$ and $CD8^+$ T cell subsets from HCs (n = 4) and patient under the indicated conditions.

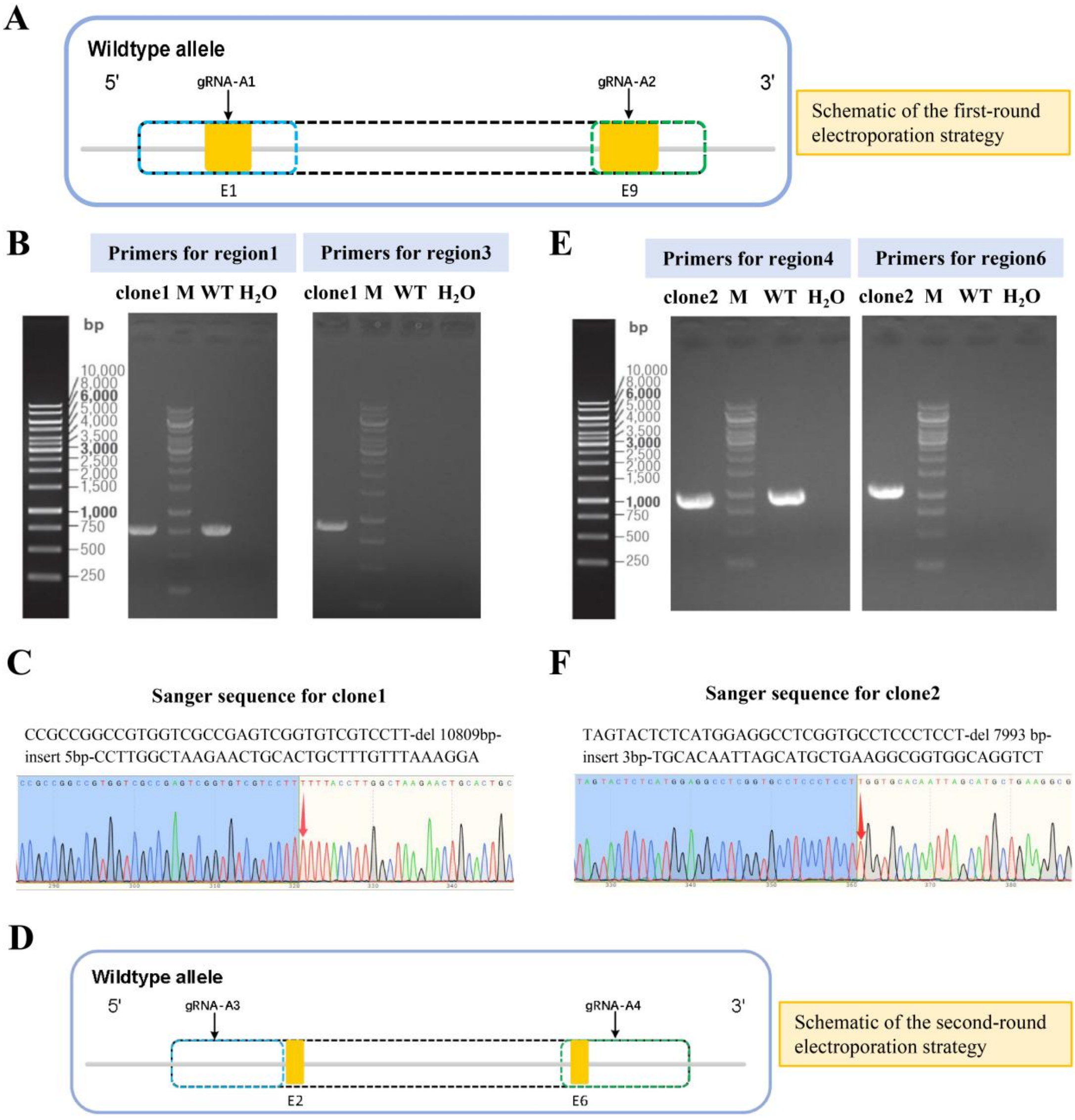


**Supplementary Figure 2. Sequential two-round CRISPR knockout strategy and validation of GTF3A in Jurkat cells.** (A) Schematic of the first-round electroporation strategy in Jurkat cells. Yellow boxes indicate the size and positions of exons; blue, green, and black boxes denote the amplification positions of Region 1, Region 2, and Region 3 primers, respectively; E, exon, e.g., E4 indicates exon 4. (B) PCR agarose gel electrophoresis of Region 1 (PCR product size: 782 bp) and Region 3 (wild-type PCR product size: 11,732 bp; Clone 1 product size: 925 bp) in Clone 1 obtained after the first round of electroporation, demonstrating heterozygosity. M, marker. (C) Sanger sequencing of Clone 1 reveals a 10,809-bp deletion and an additional 5-bp insertion at the site indicated by the red arrow. (D) Schematic of the second-round electroporation strategy in Jurkat cells. Yellow boxes indicate the size and positions of exons; blue, green, and black boxes denote the amplification positions of Region 1, Region 2, and Region 3 primers, respectively; E, exon, e.g., E4 indicates exon 4.

(E) PCR agarose gel electrophoresis of Region 4 (PCR product size: 951 bp) and Region 6 (wild-type PCR product size: 9,081 bp; Clone 1 product size: 1,088 bp) in Clone 2 obtained after the second round of electroporation. M, marker. (F) Sanger sequencing of Clone 2 reveals a 10,809-bp deletion with a 5-bp insertion, as well as a concurrent 7,993-bp deletion and a 3-bp insertion mutation in the same cell line.
